# Enhancing the Diagnostic Efficacy of Right-to-Left Shunt Using Robot-Assisted Transcranial Doppler: A Quality Improvement Project

**DOI:** 10.1101/2024.09.23.24314259

**Authors:** Ruchir Shah, Christian Devlin, Lan. Gao, Samuel Ledford, Vimal Ramjee, Vinay Madan, Jennifer Patterson, Lauren Walden, Thomas Devlin

## Abstract

**Background:** Stroke is the leading cause of adult disability worldwide, with approximately 30% of strokes remaining cryptogenic. One potential etiology is a patent foramen ovale (PFO), which may contribute to stroke through paradoxical thrombo-embolism or in situ thrombo-embolus formation. Recent advancements in robot-assisted transcranial Doppler (raTCD) have shown increased sensitivity in detecting right-to-left shunt (RLS) compared to transthoracic echocardiography (TTE), particularly in detecting large shunts, which are associated with a higher risk of stroke.

**Methods:** We conducted a retrospective quality improvement project at our regional stroke center to compare the performance of TTE and raTCD in identifying RLS in ischemic stroke patients. The study involved 148 patients admitted between February 2021 and February 2023. All patients underwent TTE and raTCD with agitated saline bubble contrast, with additional transesophageal echocardiography (TEE) at the treatment team’s discretion. The primary metrics analyzed included differences in overall RLS detection and large RLS detection rates for raTCD, TTE and TEE.

**Results:** raTCD detected RLS in 60.1% of patients compared to 37.2% with TTE (p = 0.00012), with a 42.6% detection rate for large shunts on raTCD versus 23.0% on TTE (p = 0.00053). The sensitivity and specificity of raTCD were 92% and 87.5%, respectively, compared to 78.57% and 71.43% for TTE, using TEE as the gold standard. Nine patients underwent PFO closure, all correctly identified with large shunts by raTCD, while TTE missed or underestimated the PFO size in 44% of the cases.

**Conclusion:** raTCD significantly outperforms TTE in detecting RLS and large shunts, suggesting its integration into standard PFO workup protocols may enhance secondary stroke prevention. These findings support the adoption of raTCD as a complementary diagnostic tool alongside TTE and TEE for more accurate PFO detection and risk stratification.

## INTRODUCTION

Stroke is the leading cause of adult disability in the US and worldwide and the second leading cause of adult mortality worldwide^1^. Approximately 30% of strokes remain cryptogenic^2^. Our inability to properly diagnose stroke etiology significantly jeopardizes secondary stroke preventive measures^3–6^. One potential stroke etiology is a patent foramen ovale (PFO). PFO has a reported prevalence of approximately 25% in the general population and up to 40% in patients with acute ischemic stroke^3^. It is well recognized that PFO can facilitate the passage of paradoxical thrombo-embolus from the venous to the arterial circulation or serve as a site for in situ thrombo-embolus formation. Based on the results of multiple positive PFO closure trials conducted over the past eight years, multiple American and European medical societies have published PFO diagnostic and management guidelines including indication for PFO closure^2,3,5–9^. The methodology for optimal PFO detection has been a topic of extensive investigation. A significant change in the field of PFO detection occurred with the development of robot-assisted transcranial doppler (raTCD). The results of the recently published RoBotic TCD Ultrasound BubbLe Study Compared to Transthoracic Echocardiography for Detection of Right to Left Shunt (BUBL Study – NCT04604015) reported an approximately three times higher right to left shunt rate (RLS) compared to transthoracic echocardiography (TTE)^10,11^. Furthermore, TTE completely missed or significantly downsized two thirds of large shunts detected by raTCD. These findings were deemed highly clinically relevant as PFO size is known to be a predictor of stroke risk and of central importance in calculating the risk/benefit ratio for PFO closure. Based on these results, numerous commentators have touted raTCD to be a significant medical advancement and have called for further independent “real-world” data^4,8,11–17^. To that end, we executed an independent hospital-based quality improvement project to compare the performance of TTE versus raTCD in identifying RLS in a real-world setting at a regional stroke center and assess how increased detection may translate into improved secondary stroke prevention by PFO closure.

## METHODS

This study was a single-center quality improvement project conducted at CHI Memorial Hospital in Chattanooga, TN. The study involved a retrospective chart review, with data collection occurring between February 2021 and February 2023, and chart review conducted by clinical staff from February to July 2023. The project aimed to evaluate the impact of incorporating automated transcranial Doppler (raTCD) into the stroke workflow compared to standard transthoracic echocardiography (TTE) for the diagnosis of right-to-left shunts (RLS) and patent foramen ovale (PFO). It also sought to quantitate the effect of raTCD utilization on the rate of PFO closures. The data dictionary for the study included patient demographics, raTCD results, TTE results, TEE results, and PFO closure. Local IRB waved the need for patient consent and all data was collected as part of standard clinical practice. Independent statistical analysis was funded by the non-profit NeuroScience Innovation Foundation.

### Data Collection Protocol

All imaging was collected as standard of care on patients who underwent hospitalization for an acute neurovascular episode, including ischemic stroke or transient ischemic attack. All patients underwent TTE with agitated saline bubble contrast as part of routine clinical care. Based on increasing evidence of the importance of TCD in the workup of cryptogenic stroke, raTCD was performed on all patients as part of standard cryptogenic stroke workup. All patients underwent TCD with agitated saline bubble contrast using an automated TCD platform (raTCD - NovaGuide Intelligent Ultrasound, NeuraSignal, Inc., Los Angeles, CA USA). Additional testing, such as TEE, was left to the discretion of the neurology treatment team.

The raTCD is a five degree-of-freedom robotic unit supported by artificial intelligence (AI) algorithms for the identification of the acoustic window and signal optimization using a traditional diagnostic TCD. The contrast for the TCD bubbles studies was delivered by injecting agitated saline contrast at both rest and with calibrated Valsalva, with the patient in the supine or near supine position. All raTCDs studies were performed by ultrasonographers following completion of raTCD training and demonstrated technical proficiency. The raTCD bubble studies were read by a blinded fellowship trained certified vascular neurologist and graded using the Spencer Logarithmic Scale (SLS) criteria^18^. We adopted the same definitions for a “large” RLS/PFO by raTCD as was used in the BUBL study (SLS ≥ Grade 3) and TTE RLS/PFO grading was categorized as small, moderate, or large, with reads of moderate or large being classified as large for secondary analysis^10^. All TTE bubble studies were performed by certified ultrasonographers and were read by blinded level 3 echocardiography board-certified cardiologists.

### Outcome Measures

The primary outcomes paralleled those within the BUBL study including: (a) rate of RLS detection with TTE and raTCD; (b) rate of large RLS for TTE and raTCD; (c) RLS detection rate of raTCD vs TEE; (d) RLS detection rate of TTE vs TEE; (e) sensitivity and specificity of raTCD and TTE using TEE as the gold standard; and (f) comparison of patients who received closure between raTCD and TTE for identification of RLS/PFO^10^.

### Statistical Methods

The statistical methods were performed in parallel to that performed in the original BUBL study described in detail by Rubin et al^10^. We retrospectively analyzed data from 148 ischemic stroke patients who underwent both raTCD and TTE over a 25-month period starting February 2021. Continuous variables were reported as means with standard deviations (SD) or medians with interquartile ranges (IQR), appropriately determined by normality of the data assessed using the Shapiro-Wilk test. Categorical variables were presented as frequency counts and percentages (n, %). For comparisons between categorical variables, we used chi-square tests or Fisher’s exact tests, as appropriate, to assess statistical significance. These were summarized in 2x2 tables. Two-sample proportion tests were conducted, with 95% confidence intervals and P-values reported to determine the statistical significance of the differences in detection rate of RLS/PFO between different detection tools. The exact confidence intervals were calculated using the Clopper-Pearson method for binomial proportions. Sensitivity and specificity analyses were performed using TEE as the reference standard, with corresponding confidence intervals computed using the Wilson score method without continuity correction.

All statistical analyses were performed using the statistical package R (version 4.3.1). Significance was determined at a two-tailed P-value < 0.05.

## RESULTS

We retrospectively analyzed data from 148 ischemic stroke patients who underwent both raTCD and TTE collected over a 25-month period starting February 2021. Patient demographics are presented in Table 1. Of the total population, 60.1% were male with a mean age (±S.D.) of 58.0±14.1. TTE and raTCD were performed in 100% of patients of which 14.9% (n=22) underwent TEE, TTE, and raTCD. Table 2 summarizes the overall RLS detection rate for raTCD versus TTE. Analysis revealed a 22.9% higher RLS shunt detection rate for raTCD (1.6 x higher) with a RLS shunt detection rate of 60.1% (n=89 positives) for raTCD vs 37.2% (n=55 positives) for TTE (p = 0.00012; CI: 11.2%, 34.7%). Table 3 summarizes the large RLS detection rate for raTCD versus TTE. Among the total 148 cases studied, 42.6% (n=63 positives) were identified by raTCD to have large shunts (classified as SLS Grade ≥3), representing 70% (63/89) of all positive shunts identified on raTCD. In comparison, among the total 148 cases studied, 23.0% (n=34 positives) were identified by TTE as large shunts (classified as moderate or large size on TTE), representing 61.8% (n=34/55) of all positive shunts identified by TTE, producing a difference of 19.6% (p = 0,00053; CI 8.5%, 30.7%). The agreement matrix presented in Table 4 demonstrates that of the total 9 patients in our study that underwent PFO closure 11.1% (n=1) had no PFO detected on TTE at all while no PFO that underwent closure was missed by raTCD. The agreement matrix presented in Table 5 demonstrates that all the PFOs in this study that underwent closure were designated large by raTCD; however, of those nine large/closed PFO cases, 44.4% (n=4) were misdiagnosed as being of small PFO size on TTE. The agreement matrix presented in Table 6 compares raTCD with TEE (n=33) with raTCD detecting 72.7% (24/33) and TEE 72.7% (24/33) (p = 1; CI: -21.5%, 21.1%). Table 7 shows the agreement matrix between TTE and TEE (n=22) with TTE 59.1% (13/22) and TEE detecting 63.6% (14/22) (p=1; CI: -37.8%, 28.7%). Using TEE for comparison, we calculated the sensitivity and specificity of raTCD vs TTE. Our results showed raTCD sensitivity and specificity of 95.8% and 88.9%, vs TTE sensitivity and specificity of 78.6% and 75.0%, respectively (Table 8). Of the 148 cases studied, 6.1% (n=9) had no bone window using raTCD.

**Table 1:** Patient Demographics and Testing Performed.

|  |  |
| --- | --- |
| **Total patients N** | **148** |
| **Gender, n(%)** |  |
| Female | 59 (39.9%) |
| Male | 89 (60.1%) |
| **Age** |  |
| Mean (SD) | 58.0 ± 14.1 |
| Median (IQR) | 59.0 (48.8, 68.2) |
| **Patient Cohort** |  |
| TCD performed n(%) | 148 (100.0%) |
| TTE performed n(%) | 148 (100.0%) |
| TEE performed n(%) | 22 (14.9%) |

**Table 2:**
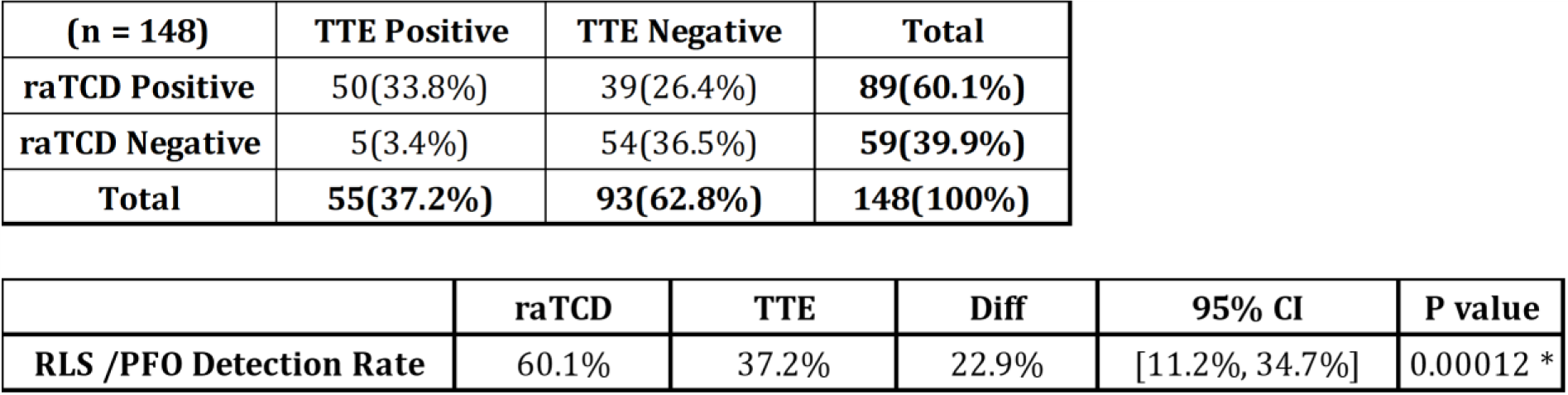
Comparison of RLS Shunt Detection for raTCD vs TTE.

**Table 3:**
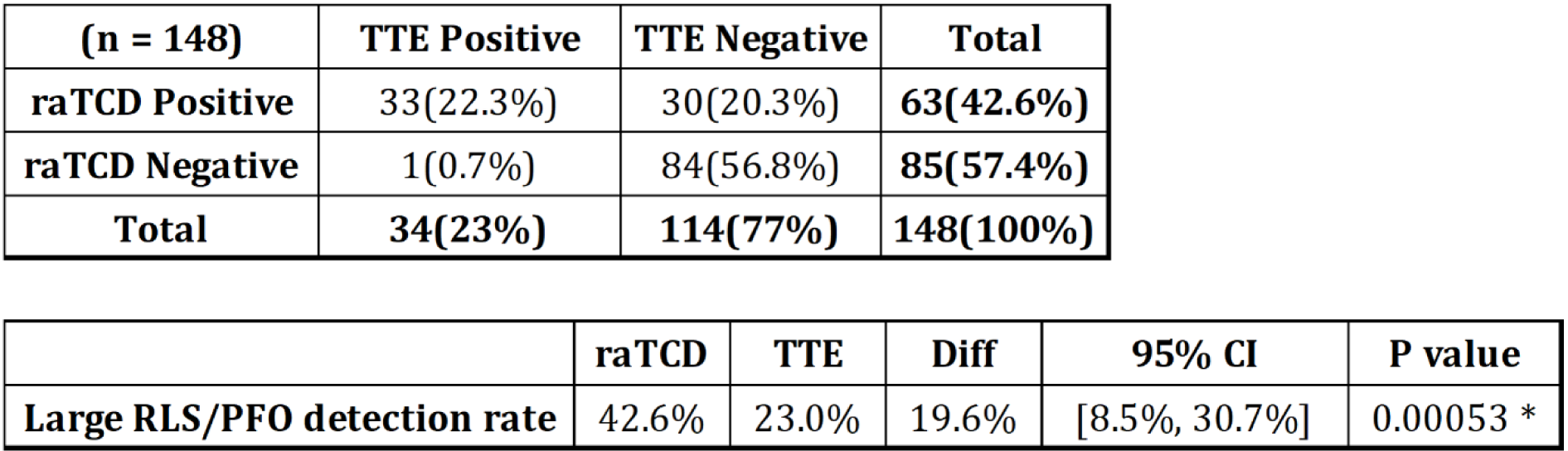
Detection rate of large RLS for raTCD vs TTE.

**Table 4:** Agreement matrix on RLS/PFO by raTCD vs TTE for PFO Closed Cases.

|  | **TTE Positive** | **TTE Negative** | **Total** |
| --- | --- | --- | --- |
| **raTCD Positive** | 8(88.9%) | 1(11.1%) | **9(100%)** |
| **raTCD Negative** | 0(0%) | 0(0%) | **0(0%)** |
| **Total** | **8(88.9%)** | **1(11.1%)** | **9(100%)** |

**Table 5:** Agreement matrix on Large RLS/PFO by raTCD and TTE for PFO Closed Cases.

|  | **TTE Large Positive** | **TTE Large Negative** | **Total** |
| --- | --- | --- | --- |
| **raTCD Large Positive** | 5(55.6%) | 4(44.4%) | **9(100%)** |
| **raTCD Large Negative** | 0(0%) | 0(0%) | **0(0%)** |
| **Total** | **5(55.6%)** | **4(44.4%)** | **9(100%)** |

**Table 6:**
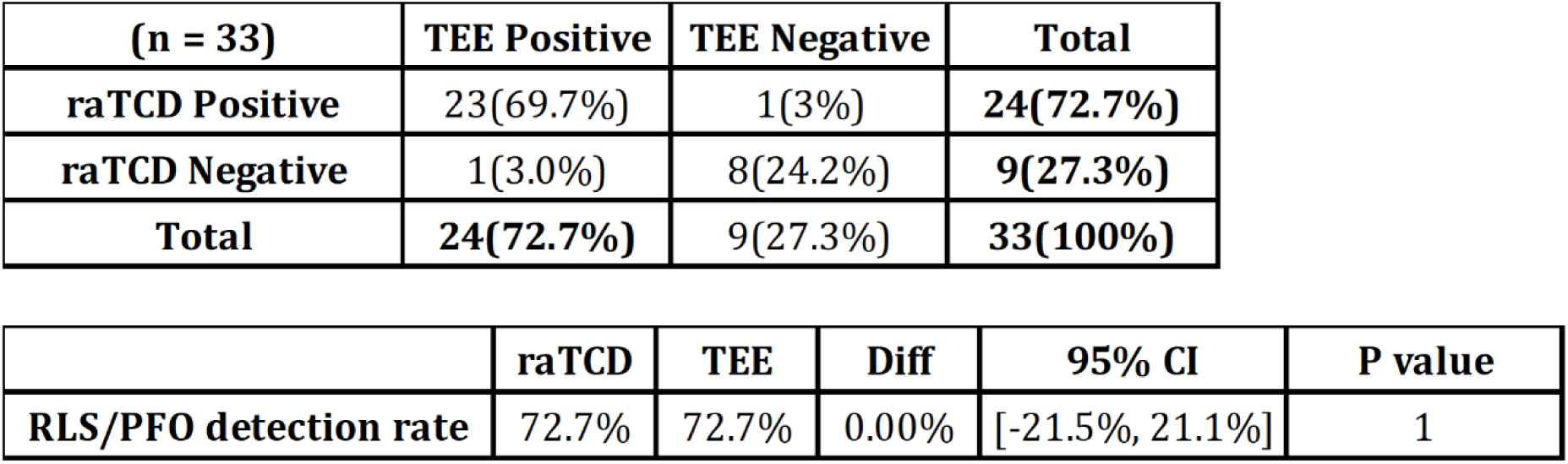
Detection rate of RLS between raTCD vs TEE for patients with TEE performed.

| **(n = 33)** | **TEE Positive** | **TEE Negative** | **Total** |
| --- | --- | --- | --- |
| **raTCD Positive** | 23(69.7%) | 1(3%) | **24(72.7%)** |
| **raTCD Negative** | 1(3.0%) | 8(24.2%) | **9(27.3%)** |
| **Total** | **24(72.7%)** | **9(27.3%)** | **33(100%)** |

|  | **raTCD** | **TEE** | **Diff** | **95% CI** | **P value** |
| --- | --- | --- | --- | --- | --- |
| **RLS/PFO detection rate** | 72.7% | 72.7% | 0.00% | [-21.5%, 21.1%] | 1 |

**Table 7:** Detection rate of RLS between TTE and TEE for patients with TEE performed.

| **(n = 22)** | **TEE Positive** | **TEE Negative** | **Total** |
| --- | --- | --- | --- |
| **TTE Positive** | 11(50.0%) | 2(9.1%) | **13(59.1%)** |
| **TTE Negative** | 3(13.6%) | 6(27.3%) | **9(40.9%)** |
| **Total** | **14(63.6%)** | **8(36.4%)** | **22(100%)** |

|  | **TTE** | **TEE** | **Diff** | **95% CI** | **P value** |
| --- | --- | --- | --- | --- | --- |
| **RLS/PFO detection rate** | 59.1% | 63.6% | -4.5% | [-37.8%, 28.7%] | 1 |

**Table 8:**
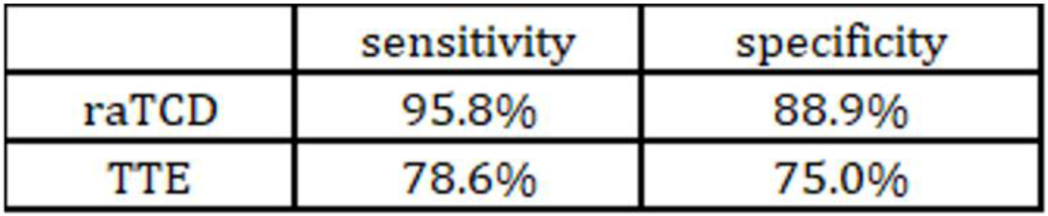
Sensitivity and Specificity for Detecting PFO (TEE as the golden standard)

|  | **sensitivity** | **specificity** |
| --- | --- | --- |
| **raTCD** | 95.8% | 88.9% |
| **TTE** | 78.6% | 75.0% |

## DISCUSSION

PFO is associated with right-to-left cardiac shunt and is a well-documented cause of embolic stroke^19^. Accurate PFO detection, sizing, and anatomic characterization are essential when assessing PFO as a potential etiology for stroke and when calculating risk/benefit ratio for PFO closure^3^. Given the multiple randomized trials showing the benefit of symptomatic PFO closure in properly selected patients, highly accurate PFO characterization is an essential step in secondary stroke prevention^3,6,9^. In this independent investigation of raTCD, reported here, we performed a retrospective review of 148 acute ischemic stroke patients admitted over a two-year period to our regional stroke center. We compared RLS detection rates and PFO size estimates based on number of bubbles detected, using raTCD and TTE in all patients and compared the findings to TEE when performed. Both the raTCD and TTE procedures followed established protocols for bubble studies, ensuring consistency and reliability in the diagnostic process. Our results corroborate the findings of Rubin et al^10^. as we found a significantly higher rate (delta = 22.9%, 1.6 x higher) of overall RLS with raTCD versus TTE (60.1% vs 37.2%, p=0.00012) and a significantly higher rate (delta = 19.6%, 1.9 x higher) of large RLS with raTCD versus TTE (42.6% vs. 23.0%, p=0.00053). Unlike Rubin et al. who did not document PFO closures, we documented that at the time of our analysis nine patients in our data set had undergone PFO closure. Our results invoke significant clinical concern as 11.1% (n=1) of patients who underwent closure had their PFO missed entirely on TTE. Furthermore, 44.4% (n=4) of the patients who underwent PFO closure had their shunts size significantly underestimated or not detected at all on TTE (all of which demonstrated large shunts on raTCD). It was owing to the performance of the raTCD that the large size of each of these cases was identified which raised the heightened alert for the need for further characterization by TEE and possible closure. The clinical importance of our findings was further underscored by the fact that 26.4% (n=39) of our study population were negative on TTE but positive on raTCD (Table 2), while only 3.4% (n=5) were detected on TTE but missed on raTCD. Similarly, we found it highly clinically concerning that 20.3% (n=30) of large RLS were missed on TTE but detected on raTCD, while only 0.7% (n=1) were missed on raTCD but detected on TTE (Table 3). Of the five subjects with a positive TTE and negative TCD, three had no windows which was known prior to the bubble contrast injection and the remaining two were negative on TEE. As TEE is often not routinely performed during the initial screen for PFO (and itself may underdiagnose PFO), the highly statistically significant improvement in our data for RLS shunt detection by raTCD over TTE suggests that many patients would be in jeopardy of having their PFO totally undiagnosed if TTE alone is used for screening. Similarly important, the highly significant difference that we report for large shunt detection rate by raTCD over TTE points to the inadequacy of TTE alone in the assessment of PFO size. Initial detection of a PFO on screening imaging may steer clinicians either towards or away from performing TEE and, as discussed below, accurate PFO sizing may be critical in calculating the risk/benefit ratio of PFO closure the ultimate driver in the close/no close decision. Therefore, reliance on TTE alone for PFO screening may lead to incorrect clinical decision making, missed opportunities for proper secondary stroke prevention by PFO closure, and increased subsequent stroke risk for patients. Finally, our study was able to expand on Rubin et al. by reporting sensitivity and specificity for both raTCD and TTE compared to TEE (sensitivity of 95.8% and specificity of 88.9% for raTCD and sensitivity of 78.6% and specificity of 75.0% for TTE). While our overall total RLS detection rate by raTCD (60.1%) was almost identical to that reported by Rubin et al. (64%), our overall rate of positive RLS detected by TTE was higher (37.2% vs 20%). When compared to the report by Rubin et al., we found a substantially higher rate of large RLS on raTCD (42.6% vs 27.0%) and on TTE (23.0% vs 10%). The reason for our higher detection rates of RLS on raTCD for larger PFOs and for all PFOs on TTE is uncertain. This variability between our single center results and the multi-center trial reported by Rubin may have involved multiple factors discussed in detail below.

### Concordance of Studies Supporting a Central Role for TCD in RLS Detection

Numerous investigations have compared various methodology for PFO detection including TTE, TEE, and TCD^13,15–17,20–23^. The development of raTCD has emerged as a potentially important advancement in the field of PFO detection. The recent muti-center investigation comparing raTCD versus TTE by Rubin et al. reported a 41.4% overall higher rate of RLS detection in stroke patients and a 17.4% higher detection of large RLS by raTCD over TTE^10^. The accompanying editorial underscored the potential significance of Rubin’s findings and called for additional investigations comparing raTCD to TTE and TEE^11^. Similarly, Rubin’s findings led to a direct call for further investigation of raTCD by the Roundtable of Academia and Industry for Stroke Prevention (RAISE)^3^. Based on the current literature, RAISE endorsed raTCD being considered in the current workup for PFO^3^. In a recent commentary by Dr. Braydon Dymm titled “Could Robot-Assisted Transcranial Dopler Replace Transthoracic Echocardiography as Screening for Right to Left Shunt After Cryptogenic Stroke” they challenged the current standard of TTE per se as the initial screening tool for PFO detection in patients with cryptogenic stroke^24^.

Our results are consistent with previous reports comparing TTE, TCD, and TEE for the detection of PFO. Mojadidi et al. in 2014 published a meta-analysis of 27 studies (29 comparisons) and 1,968 patients with the aim of determining the accuracy of TCD using TEE as a reference. Their meta-analysis showed that TCD had a sensitivity, specificity, positive likelihood ratio (LR+), and negative likelihood ratio (LR-) of 97%, 93%, 14, and 0.04, respectively. These authors concluded that TCD is the preferred test for detecting RLS in patients with cryptogenic stroke or migraine^25^. A more recent meta-analysis (35 studies, 3,067 patients) reported a sensitivity, specificity, and area-under-cure for PFO diagnosis by TCD of 96.1%, 92.4%, and 0.98 versus 45.1%, 99.6%, and 0.86 for TTE. The analysis also reported LR+ and LR- for the TCD (12.62, 0.04) and TTE (106.61, 0.55) respectively leading to their conclusion of the diagnostic superiority of TCD in comparison to TTE for RLS detection^21^. A more recent study (775 patients with TCD and TTE) reported significant test superiority (dominance) of TCD over TTE for RLS/PFO detection and concluded with the recommendation for TCD as the preferred screening method for RLS despite the limitations of differentiating between intracardiac and extracardiac shunts^26^. Similar results were also reported in a 2018 investigation suggesting the need for TCD within the PFO work up^27^. This recent widespread interest in TCD is in line with the joint European position paper involving nine separate national societies which in 2019 called for TCD to be incorporated as an essential part of PFO workup^8^. Similarly, the European Stroke Organization (ESO) in their 2024 published an expert consensus statement that “as there is no technique that can be considered as a gold standard, we advise locally agreed diagnostic algorithms using the available techniques (TCD, TTE and TOE) to diagnose an RLS”. In that statement they cited the recent Rubin et al. study as support^28^. These various publications are summarized in Table 9 comparing the overall performance of TCD, TTE, and TEE.

**Table 9:**
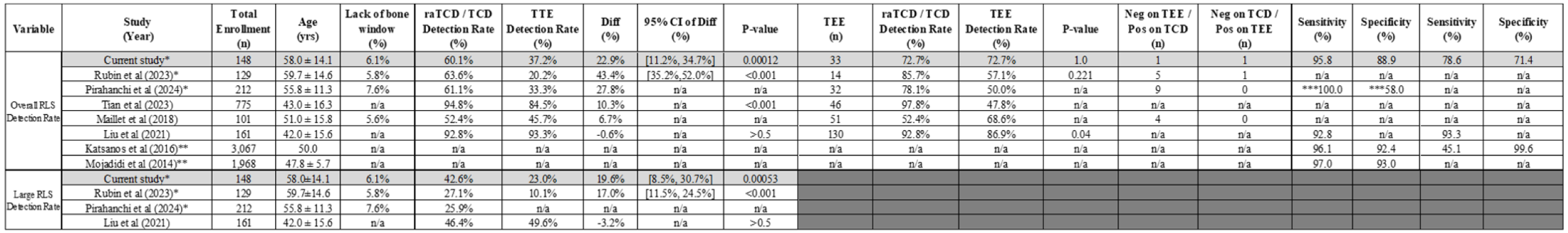
Comparison of detection rate of raTCD/TCD vs TTE and TEE between various other published studies.

### Unique Aspects of raTCD That May Lead to Increased Accuracy of raTCD

The major limitation of TCD is the inability to distinguish between non-cardiac and cardiac RLS. Despite the fact that PFO is the most likely diagnosis in RLS if bubbles are detected within 3-5 cardiac cycles of injection, current guidelines recommend that all possible PFO’s detected by TCD be evaluated by TEE prior to attempted closure^3^. This is critical to confirm the absence of an extracardiac shunt and for anatomic PFO characterization. Despite this limitation, TCD has specific advantages in the assessment of RLS/PFO including: 1) a high bubble detection rate through the intracranial circulation in the very narrow diameter middle cerebral artery whose directionality of bubbles are all aligned with the angle of insonation; 2) the ability to quantify and confirm an adequate Valsalva maneuver by measuring a ≥ 25% drop in MCA flow velocities; and 3) unlike TTE and TEE, TCD is typically performed with the patient in a semi-upright position which has been demonstrated to be associated with higher rates of PFO detection. Furthermore, there appear to be additional technical advantages of raTCD that increase its ability to detect RLS. These include 1) bilateral simultaneous automated insonation with vessel imaging optimization (alleviating the need or a trained vascular technologist); 2) raTCD incorporates advanced software to aid the user in identification of the regions of interest for the study as well as full audio and video playback to aid in distinguishing bubbles injected peripherally from artifact; 3) unlike TTE (in which atrial images are partially obscured by the ribs) and manual TCD (which has a higher rate of unsuccessful insonation due to poor bone windows), raTCD offers excellent visualization of the unobstructed intracranial vasculature for bubble detection^29^ including “no window” rates of 6.1% in the current study, 5.8% in BUBL study by Rubin et al^10^ and 3.5% in a group of healthy volunteers^30^; and 4) utilization of Power M-Mode TCD which has been shown to be more sensitive for RLS detection compared to single gated manual TCD^18^.

### Impact of study protocol on RLS screening test accuracy

In addition to technological differences between imaging modalities, protocol specifics can have a dramatic impact on the accuracy of tests. Valsalva maneuver appears to play a major role in the ability to detect a positive bubble study on TEE or TTE. Caputi et al. reported an overall PFO detection rate by TEE of 63% with a general concordance between manual TCD and TEE of approximately 90% with a sensitivity and specificity for cTCD of 96.8% and 78.4% respectively^13^. Without Valsalva, TCD was far superior at overall RLS detection (75%) over TEE (48%)(p<0.001). Furthermore, of the patients that demonstrated a “shower-curtain” pattern on TCD at rest (indicating very large RLS), only 71% of those patients exhibited RLS at all on TEE at rest. Of patients that exhibited a smaller, “non-curtain effect”, RLS at rest on TCD, only 22% of those were noted to have any RLS on TEE at rest. These findings indicate the highly operator dependent nature of TEE and the critical aspect of Valsalva when performing TEE and TCD. Based on their results and the individual benefits of both TEE and TCD, Caputi et al. proposed that “the combination of TEE and TCD could be considered the real gold standard for PFO”^13^. The critical importance of Valsalva also applies to TTE where the maneuver has been reported to increase specificity from 40% to 60%^14^. While adequate Valsalva can be easily confirmed quantitatively on raTCD by a drop in the MCA flow velocity of approximately 25%, adequate Valsalva on TTE or TEE is much harder to confirm and in the case of TEE harder to elicit due to patient sedation. Difficulty with patient tolerability of the TEE probe, variations in cardiac anatomy and operator experience all may contribute to reducing the sensitivity of TTE and TEE in PFO detection. Patient positioning also appears to be an additional potentially important factor in PFO detection. Lucreziotti et al. reported increased RLS on TCD and TTE with patients in the sitting position compared to the supine position^29^. Regarding raTCD, Rubin et al. reported similar results in their secondary analysis of the BUBL study showing that both Valsalva and bed positioning (HOB angle 0° to 45°) had a significant impact on RLS grade whereas IV location did not^31^. As TTE is typically performed with the patient semi-prone, this may be a driver to lower performance. In our study however (where raTCD was performed at or below 45° incline) it is unlikely that this factor is a major contributing factor to our results. Given the known positional effects on PFO detection it would be theoretically ideal to compare raTCD and TTE simultaneously in identical positions. Practically speaking, however, optimal imaging windows on TTE are highly positional dependent based on body habitus and are often suboptimal in the semi-erect position.

Despite strong support in the literature for the role of TCD when performed in conjunction to TTE for RLS detection, variability does exist in the reported specificity of TTE for RLS detection (see Table 9). A paper published in Echocardiography in 2020 (161 patients with various neurological disorders) utilizing right heart catheterization as the gold standard reported a sensitivity of 92.75% for TCD and 93.33% for TTE, but specificity was not reported^32^. TTE image quality is known to be highly dependent on BMI^33^. Therefore, well-established regional differences in BMI nationally (highest in our region of the south) may contribute to the variability in published RLS detection rates using TTE^34^. In addition to a multitude of other variables (ex., ultasonographer experience, degree of patient hydration, hardware and software variability, etc.), Valsalva technique is a critical determinant of RLS detection rates on TTE^35^. As most centers, including ours, do not use calibrated Valsalva during standard TTE bubble studies, variation in Valsalva may contribute significantly to the variability in reported rates of RLS on TTE. The ability of raTCD to mitigate the magnitude of the influence of many of these variables appears advantageous^13,14,20,21^.

### Rational for the New Protocol for PFO Workup Incorporating raTCD

Recommendations are emerging in numerous review articles and society guidelines in the United States and from Europe, that the concept of TEE as a “gold standard” be replaced by the concept that TEE, TCD, and TTE each have their own unique strengths and limitations and that proper PFO screening requires a coordinated protocol that often involves all three. Based on our results and the numerous other investigations discussed above, we now propose the following protocol as shown schematically in Figure 1. This protocol builds upon that presented in the muti-society European guidelines published in 2019 and the recent ESO guidelines on the diagnosis and management of PFO ^8,28^. TTE with agitated saline injection is the logical initial diagnostic test to be performed to screen for PFO. Given its non-invasive nature and the need to assess for other causes of embolic source (mural wall thrombus, low ejection fraction, wall motion abnormalities, or significant valvular abnormalities), TTE with bubble study is the obvious best initial screening tool. Next, if no PFO or a small PFO is detected on TTE, TCD should then be performed. We favor raTCD based on its advantages listed above and it significant increased RLS detection rate compared with manual TCD^10^. The benefit of raTCD stems from its increased sensitivity for overall RLS detection and its superior ability to identify a larger number of bubbles leading to a more accurate estimate of shunt size. While numerous scales, such as the ROPE score, have been employed to assess likelihood of PFO as a causative factor for stroke, more refined scales (ex., PASCAL) now incorporate PFO size and the presence of high-risk structural features (atrial septal aneurysm (ASA))^36,37^. PASCAL allows for an easy and highly clinically useful validated calculation of the risk/benefit ratio of PFO closure^3^. When calculating a PASCAL score to determine risk/benefit ratio for potential PFO closure on a patient with cryptogenic stroke, the clinician must go through a 4-step process of: 1) accurately identifying that a PFO is present; 2) calculating the ROPE score based on the patients history; 3) estimating the size of the PFO (if PFO is absent or small on TTE, raTCD should be performed); and 4) utilization of TEE to rule out a non-cardiac cause of RLS and determining the presence or absence of an associated ASA. With this data in hand, the PASCAL score can then be calculated^3^. Based on PASCAL, if the PFO is determined to be the “possible” or “probable” stroke etiology, the risk/benefit ratio may favor PFO closure in properly selected patients. If PASCAL determines that the PFO is an “unlikely” etiology of stroke, PFO closure may be associated with increased harm. Even in the absence of an ASA, the identification of a larger PFO (which may be detected on raTCD but undersized or missed on TTE or TEE) in the setting of a high ROPE scale score may result in a moderate risk PASCAL PFO score qualifying for closure^3^. Therefore, missing a PFO completely or mislabeling a PFO as small based on erroneous TTE/TEE information may lead to loss of opportunity for proper secondary stroke prevention by PFO closure. Similarly, the presence of ASA, even in the presence of a small PFO, combined with high ROPE score may qualify as a “possible” risk PFO on PASCAL thereby justifying PFO closure. For these reasons, a stepwise protocol utilizing TTE, raTCD, and TEE appears to be the best approach for PFO detection and rigorous adjudication to properly determine the risk/benefit ratio for PFO closure.

**Figure 1:**
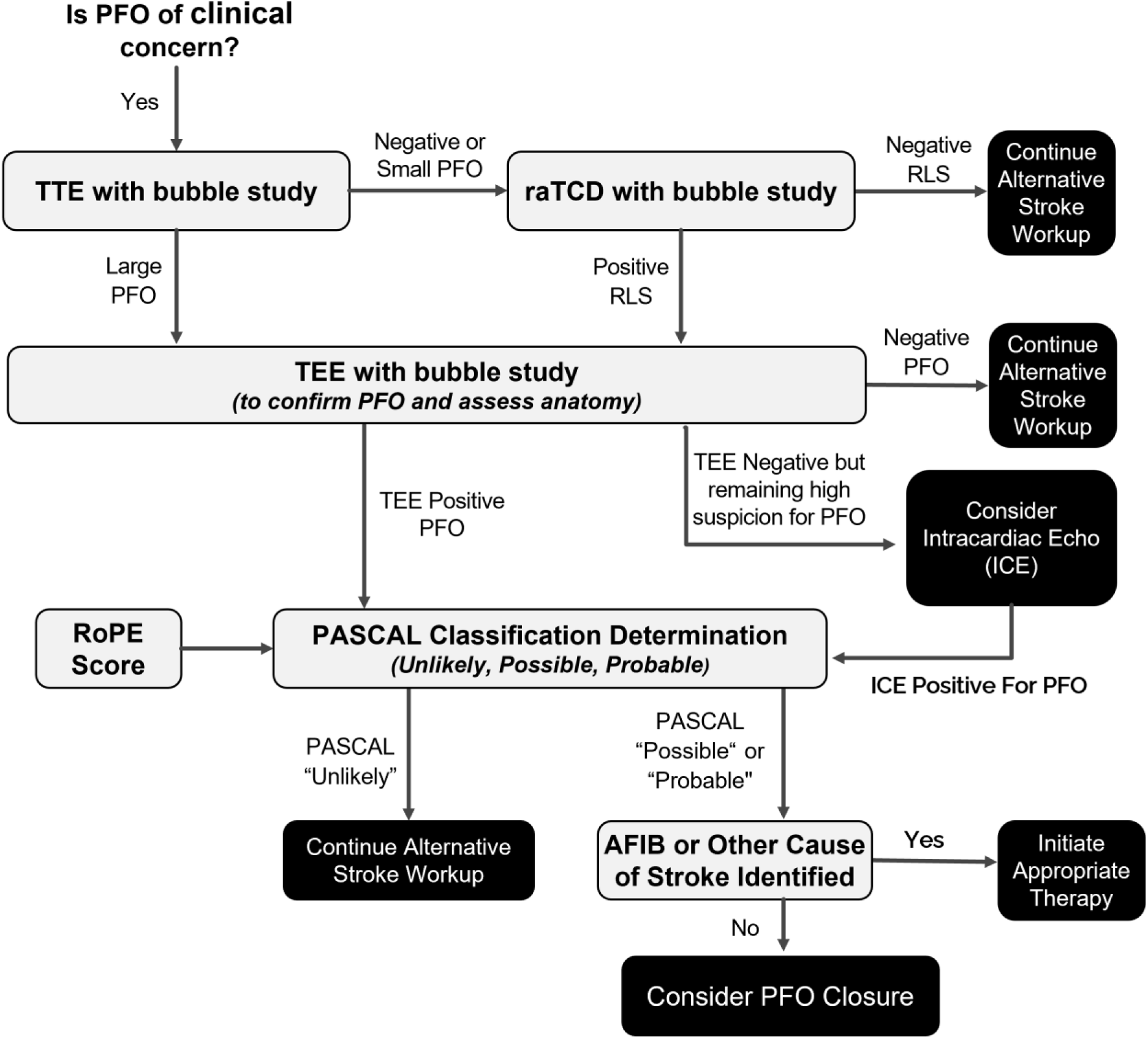
Recommended Diagnostic Algorithm for Patent Foramen Ovale (PFO) Diagnosis and Patient Selection for PFO Closure.

The preponderance of evidence discussed above now indicates that by utilizing a combination of TTE, TEE, and raTCD, as outlined in Figure 1, we can significantly reduce the likelihood of missed PFO diagnoses that could otherwise occur with the exclusive use of TTE and/or TEE. Recent publications also suggest that in rare cases in which a high index of suspicion for PFO remains (ex., positive raTCD but negative TTE/TEE), intracardiac echocardiography (ICE) or other invasive intracardiac imaging can be considered^38–40^. Ongoing large-scale comparisons of TCD, TTE and TEE performance and studies on how best to optimize each should continue to be reported. The cumulative literature suggests that the etiology of stroke for countless patients has likely gone shrouded in mystery for years. It is high time to define a new standard of care approach for PFO screening based on a composite assessment with TTE-TEE-raTCD to fulfill our commitment to our patients to provide the highest-level secondary stroke prevention.

## Data Availability

All data related to this publication are available on request to the corresponding author.

## REFERENCES

1. Katan M, Luft A. Global Burden of Stroke. Semin Neurol. 2018;38(02):208–211.

2. Prabhakaran S, Messé SR, Kleindorfer D, et al. Cryptogenic stroke: Contemporary trends, treatments, and outcomes in the United States. Neurol Clin Pract. 2020;10(5):396–405.

3. Sposato LA, Albin CSW, Elkind MSV, et al. Patent Foramen Ovale Management for Secondary Stroke Prevention: State-of-the-Art Appraisal of Current Evidence. Stroke. 2024;55(1):236–247.

4. Katsanos, et al. A. TCD VS TTE for PFO - REVIEW. Am Neurol Assoc. 2016;79(4):625–635.

5. Patel U, Dengri C, Pielykh D, et al. Secondary Prevention of Cryptogenic Stroke and Outcomes Following Surgical Patent Foramen Ovale Closure Plus Medical Therapy vs. Medical Therapy Alone: An Umbrella Meta-Analysis of Eight Meta-Analyses Covering Seventeen Countries. Cardiol Res. 2023;14(5):342–350.

6. Safouris A, Kargiotis O, Psychogios K, et al. A Narrative and Critical Review of Randomized-Controlled Clinical Trials on Patent Foramen Ovale Closure for Reducing the Risk of Stroke Recurrence. Front Neurol. 2020;11:434.

7. Kleindorfer DO, Towfighi A, Chaturvedi S, et al. 2021 Guideline for the Prevention of Stroke in Patients With Stroke and Transient Ischemic Attack: A Guideline From the American Heart Association/American Stroke Association. Stroke. 2021;52(7).

8. Pristipino C, Sievert H, D’Ascenzo F, et al. European position paper on the management of patients with patent foramen ovale. General approach and left circulation thromboembolism. Eur Heart J. 2019;40(38):3182–3195.

9. Shi F, Sha L, Li H, et al. Recent progress in patent foramen ovale and related neurological diseases: A narrative review. Front Neurol. 2023;14:1129062.

10. Rubin MN, Shah R, Devlin T, et al. Robot-Assisted Transcranial Doppler Versus Transthoracic Echocardiography for Right to Left Shunt Detection. Stroke. 2023;54(11):2842–2850.

11. Wechsler LR. Robot-Assisted TCD for Detection of Right to Left Shunt: Teaching an Old Device New Tricks. Stroke. 2023;54(11):2851–2852.

12. Grisold A, Rinner W, Paul A, et al. Estimation of patent foramen ovale size using transcranial Doppler ultrasound in patients with ischemic stroke. J Neuroimaging. 2022;32(1):97–103.

13. Caputi L, Carriero M, Falcone C, Oiotti P, et al. Transcranial Doppler and Transesophageal Echocardiography: Comparison of Both Techniques and Prospective Clinical RElevance of Transcranial Doppler in Patent Foramen Ovale Detection. J Stroke Cerebrovasc Dis. 2009;18(5):343–348.

14. Nemec JJ, Marwick TH, Lorig RJ, et al. Comparison of transcranial Doppler ultrasound and transesophageal contrast echocardiography in the detection of interatrial right-to-left shunts. Am J Cardiol. 1991;68(15):1498–1502.

15. Mojadidi. Diagnostic Accuracy of Transesophageal Echocardiogram for the Detection of Patent Foramen Ovale: A Meta-Analysis. Echocardiography. 2014;31(6):752–758.

16. Pirahanchi Y, Isaguirre S, Rodriguez-Brizuela R. Incorporation of Autoated Robotic Transcranial Doppler to Screen for PFO and Qualntify R to L Shunt Severity in Evaluation of Ischmic Stroke Patients for Stroke Etiology at a Comprehensive Stroke Center. Stroke. 55(Suppl 1).

17. Hutayanon P, Muengtaweepongsa S. The Role of Transcranial Doppler in Detecting Patent Foramen Ovale. J Vasc Ultrasound. 2023;47(1):33–39.

18. Spencer,Merrill, Moehring M, Jesurum J, Gray W, Olsen J. Power M-Mode Transcranial Doppler for Diagnosis of Patent Foramen Ovale and Assessing Transcatheter Closure. J Neuroimaging. 2006;14(4):342–349.

19. Saposnik G, Bushnell C, Coutinho JM, et al. Diagnosis and Management of Cerebral Venous Thrombosis: A Scientific Statement From the American Heart Association. Stroke. 2024;55(3).

20. González-Alujas T, Evangelista A, Santamarina E, et al. Diagnosis and Quantification of Patent Foramen Ovale. Which Is the Reference Technique? Simultaneous Study With Transcranial Doppler, Transthoracic and Transesophageal Echocardiography. Rev Esp Cardiol Engl Ed. 2011;64(2):133-139.

21. Katsanos, et al. A. Transcranial Doppler versus transthoracic echocardiography for the detection of patent foramen ovale in patients with cryptogenic cerebral ischemia: A systematic review and diagnostic test accuracy meta-analysis. Ann Neurol. 2016;79(4):625–635.

22. Van Der Giessen H, Wilson LC, Coffey S, Whalley GA. Review: Detection of patient foramen ovale using transcranial Doppler or standard echocardiography. Australas J Ultrasound Med. 2020;23(4):210–219.

23. Palazzo P, Ingrand P, Agius P, Belhadj Chaidi R, Neau J. Transcranial Doppler to detect right-to-left shunt in cryptogenic acute ischemic stroke. Brain Behav. 2019;9(1):e01091.

24. Rubin M, Shah R, Devlin T, et al. Commentary: Could Robot-Assisted Transcranial Doppler Replace Transthoracic Echocardiography as Screening for Right to Left Shunt After Cryptogenic Stroke? Blogging Stroke. Published online February 5, 2024. Accessed July 12, 2024.

25. Mojadidi MK, Roberts SC, Winoker JS, et al. Accuracy of Transcranial Doppler for the Diagnosis of Intracardiac Right-to-Left Shunt. JACC Cardiovasc Imaging. 2014;7(3):236–250.

26. Tian L, Zhang M, Nie H, Zhang G, Luo X, Yuan H. Contrast-enhanced transcranial doppler versus contrast transthoracic echocardiography for right-to-left shunt diagnosis. J Clin Monit Comput. 2023;37(5):1145–1151.

27. Maillet A, Pavero A, Salaun P, et al. Transcranial Doppler to Detect Right to left Communication: Evaluation Versus Transesophageal Echocardiography in Real Life. Angiology. 2018;69(1):79–82.

28. Caso V, Turc G, Pristipino C. European Stroke Organisation (ESO) Guidelines on the diagnosis and management of patent foramen ovale (PFO) after stroke. Eur Stroke J. Published online May 16, 2024.

29. Lucreziotti S, Debenedetti C, Massironi L, Mantero A. The effect of posture in patients with patent foramen ovale: Evaluation of the right-to-left shunt with transcranial Doppler and transthoracic echocardiography and correlation with the arterial oxygen saturation. G Ital Cardiol. 2017;18(6):519–524.

30. O’Brien MJ, Dorn AY, Ranjbaran M, et al. Fully Automated Transcranial Doppler Ultrasound for Middle Cerebral Artery Insonation. J Neurosonology Neuroimaging. 2022;14(1):27–34.

31. Rubin M, Shah R, Youn T, et al. Transcranial Doppler Bubble Study Technique in a Clinical Trial: Valsalva Maneuver and Body Position Matter but IV Location Does Not (S10.002). Neurology. 2024;102.

32. Liu F, Kong Q, Zhang X, et al. Comparative analysis of the diagnostic value of several methods for the diagnosis of patent foamen ovale. Echocardiography. 2021;(38):790–797.

33. Ellenberger K, Jeyaprakash P, Sivapathan S, et al. The Effect of Obesity on Echocardiographic Image Quality. Heart Lung Circ. 2022;31(2):207–215.

34. Myers CA, Slack T, Martin CK, Broyles ST, Heymsfield SB. Regional disparities in obesity prevalence in the United States: A spatial regime analysis: Regional Disparities in Adult Obesity. Obesity. 2015;23(2):481–487.

35. Stafford MB, Bagley JE, DiGiacinto D. Comparison of Transthoracic Echocardiography, Transesophageal Echocardiography, and Transcranial Doppler in the Detection of Patent Foramen Ovale as the Etiology for Cryptogenic Stroke. J Diagn Med Sonogr. 2019;35(2):127–133.

36. Kent DM, Saver JL, Kasner SE, et al. Heterogeneity of Treatment Effects in an Analysis of Pooled Individual Patient Data From Randomized Trials of Device Closure of Patent Foramen Ovale After Stroke. JAMA. 2021;326(22):2277.

37. Lee OH, Kim JS. Percutaneous Patent Foramen Ovale Closure After Stroke. Korean Circ J. 2022;52(11):801.

38. Han KN, Yang SW, Zhou YJ. Novel way of patent foramen ovale detection and percutaneous closure by intracardiac echocardiography: A case report. World J Clin Cases. 2022;10(29):10559–10564.

39. Han KN, Ma XT, Yang SW, Zhou YJ. Intracardiac echocardiography in the diagnosis and closure of patent foramen ovale. J Geriatr Cardiol. 2021;18(9): 697.

40. Peters A, Pravin P. TEE Versus ICE in Structural Interventions - American College of Cardiology. Am Coll Cardiol. Published online September 10, 2018. https://www.acc.org/Latest-in-Cardiology/Articles/2018/09/10/08/09/TEE-Versus-ICE-in-Structural-Interventions

